# Community Transmission of SARS-CoV-2 by Fomites: Risks and Risk Reduction Strategies

**DOI:** 10.1101/2020.11.20.20220749

**Authors:** Ana K. Pitol, Timothy R. Julian

## Abstract

SARS-CoV-2, the virus responsible for the COVID-19 pandemic, is perceived to be primarily transmitted via person-to-person contact, through droplets produced while talking, coughing, and sneezing. Transmission may also occur through other routes, including contaminated surfaces; nevertheless, the role that surfaces have on the spread of the disease remains contested. Here we use the Quantitative Microbial Risk Assessment framework to examine the risks of community transmission of SARS-CoV-2 through contaminated surfaces and to evaluate the effectiveness of hand and surface disinfection as potential interventions. The risks posed by contacting surfaces in communities are low (average of the median risks 1.6×10^−4^ - 5.6×10^−9^) for community infection prevalence rates ranging from 0.2-5%. Hand disinfection substantially reduces relative risks of transmission independently of the disease’s prevalence and the frequency of contact, even with low (25% of people) or moderate (50% of people) compliance. In contrast, the effectiveness of surface disinfection is highly dependent on the prevalence and the frequency of contacts. The work supports the current perception that contaminated surfaces are not a primary mode of transmission of SARS-CoV-2 and affirms the benefits of making hand disinfectants widely available.

## Introduction

SARS-CoV-2, the virus responsible for the COVID-19 pandemic, is transmitted primarily via person-to-person pathways such as prolonged exposures to respiratory droplets produced while talking, coughing, and sneezing ^1,2^. Based on the assumption of respiratory-droplet transmission, infection control recommendations include maintaining social/physical distances, wearing masks, case isolation, contact tracing, and quarantine ^3^. Due to the possibility of transmission through other routes, including airborne and surface-mediated transmission, the WHO recommends taking airborne precautions for particular settings where aerosols are generated and emphasizes the importance of hand hygiene^2^. Nevertheless, the role that airborne and surface-mediated transmission have on the spread of the disease remains contested ^1,2,4–7^.

Indirect transmission via fomites (contaminated surfaces) contributes to the spread of common respiratory pathogens ^8–10^ and evidence-to-date suggests fomite transmission is possible for SARS-CoV-2. People infected with SARS-CoV-2 shed the virus into the environment, as evidenced by extensive SARS-CoV-2 RNA detected on surfaces in cruise ships, hospitals, and public spaces in urban areas such as bus stations and public squares ^11–15^. Infective coronavirus persists in the environment, with experimental evidence of persistence on surfaces ranging from 3 hours to 28 days, depending on environmental factors such as surface material and temperature ^16–18^. Viruses readily transfer from contaminated surfaces to the hand upon contact ^19–21^ and from hands to the mucous membranes on the face ^21–23^. People touch their faces frequently, with studies reporting average hand-to-face contacts ranging from 16 to 37 times an hour ^24–26^. Taken together, this suggests surface contamination could pose a risk for indirect SARS-CoV-2 transmission, similar to other respiratory viruses^8^.

Despite the potential importance of indirect transmission, it is difficult to estimate its role relative to direct transmission. Quantitative Microbial Risk Analysis (QMRA) provides a framework for understanding health risks from indirect transmission and provides insights into potential impacts of infection control recommendations. Mechanistic models of transmission events within the context of QMRA frameworks have been used to identify risks for a number of scenarios including children playing with fomites ^27^, sanitation workers collecting and processing urine for nutrient recovery ^28^, and people sharing a houseboat.^29^ Within the context of the current COVID-19 pandemic, QMRA has been adapted to evaluate and compare transmission risks for MERS-CoV and SARS-CoV-2 through droplets, aerosolized particles, and doffing personal protective equipment in hospitals ^30–32^ and to evaluate the effectiveness masks at reducing the risk of SARS-CoV-2 infection ^33^.

In this study, two mechanistic models of indirect transmission within the QMRA framework are used to estimate the risk of infection for SARS-CoV-2 in community settings and inform guidance on potential intervention strategies. Specifically, a model is developed to estimate the risk of infection for single contacts with contaminated surfaces, with the concentrations of SARS-CoV-2 RNA on the surfaces informed by literature investigating surface contamination in public spaces (bus stations, gas stations, stores, playgrounds). A second model is used to estimate risks from surface-mediated community transmission as a function of the prevalence of COVID-19 cases in the community and to test the efficacy of feasible intervention strategies of hand disinfection and surface disinfection.

## Methods

### Model 1. Risks from contaminated surfaces

A stochastic-mechanistic model was developed to estimate the infection risk for a single hand-to-surface followed by hand-to-face contact (Figure S1). The concentration of SARS-CoV-2 RNA on public surfaces [gene copy number (gc) cm^-2^] was obtained from literature^13,15^. Conversion of SARS-CoV-2 RNA to infective virus was assumed to follow a uniform distribution with range 100 and 1000 (gc per infective virus, with infective virus measured using Plaque Forming Units (PFU)). The gc:PFU ratio is based on the sparsely available information of SARS-CoV-2 found in literature ^18,34,35^, data from enveloped respiratory RNA viruses^36^ (seasonal influenza A(H1N1), A(H3N2), and influenza B have mean ratios of 708, 547, and 185 gene copies per TCID_50_ respectively), and a ratio of 0.7 to convert TCID_50_ to PFU^37^. The transfer of virus from surface-to-hand and from hand-to-mucous membranes was assumed to be proportional to the concentration of virus on the surface and the transfer efficiency of virus at both interfaces^38^. An exponential dose-response model^39^ was used to estimate the probability of infection for a given dose. This model is based on the pooled data of studies of SARS-CoV^40^ and Murine hepatitis virus (MHV-1)^41^ infection in mice. The upper bounds of the dose-response curve are consistent with the infectivity of two different variants of SARS-CoV-2 in mice, hamsters, and ferrets^42^. Monte Carlo simulations were used to incorporate the uncertainty and variability of the input parameters. The model was simulated 50,000 times. Results are presented as the median risk values with 5^th^ and 95^th^ percentiles. The equations used, the probability distributions for the input parameters, and a diagram of the model can be found in the Supporting Information (Figure S1-S3, Table S1).

### Model 2. Risks from surface-mediated community transmission

Contamination of SARS-CoV-2 on surfaces in public spaces (e.g., traffic light buttons, train buttons) was modeled as a function of disease prevalence in the community and frequency of contact with the surface. Estimates obtained in the model describe the probability of infection for people contacting the surface across a period of seven days. In the model, surface inoculation happens when infected individuals use their hand to cover their mouth while coughing and subsequently touching a surface. Viral loads [gc mL^-1^] in the saliva or sputum of symptomatic COVID-19 patients within the first 14 days of symptom onset were used as input to the model^35,43–45^. The concentrations of SARS-CoV-2 in saliva and sputum samples measured in genome copies35,43–45, align with concentrations of samples measured in TCID_50_ ^34^ once they are adjusted by the previously mentioned genome copies to infectivity ratio.

The frequency of surface contamination was determined by the prevalence of the disease in the population^46–50^. A cough was assumed to spread particles conically^51^. Therefore, virus inoculation on hands was modeled as a function of the concentration of virus in the saliva, the volume of saliva expelled per cough, the distance between the mouth and the hand, and the right circular cone angle of the ejected particles, *a* (Figure S2, Table S1). Transfer from surface-to-hand and from hand-to-mucous membranes was assumed proportional to the concentration of virus on the surface and the transfer efficiency of virus at both interfaces^38^ (Figure S1). The concentration of virus in the contaminated surface was assumed to decay exponentially ^52^. Decay rate was obtained from research on SARS-CoV-2 survival on various surfaces ^18^. An exponential dose-response model^39^ was used to estimate the probability of infection for a given dose. The concentration on the contaminated surface and on the hand was reduced according to the log_10_ reduction values for the scenarios of surface and hand disinfection. Alcohol-based hand sanitizer was selected as hand disinfection strategy due to the widespread availability and portability of hand-sanitizers. Although hand-washing was not considered, based on the log reductions of enveloped viruses achieved by handwashing^53^, we assume effectiveness of handwashing is similar to hand sanitizer for the reduction of SARS-CoV-2 on hands.

Monte Carlo simulations were used to incorporate uncertainty and variability of the input parameters in the risk characterization. Convergence was tested for the baseline scenario by running five times 5 000, 10 000, 20 000, 50 000, and 100 000 simulations. There was minimal variation after 50 000 simulations (Supplementary Figure 2). Based on the results, all the models were simulated 50,000 times. For each of the 50,000 simulations, the risks were calculated across time, for a period of seven days. Therefore, each simulation has a time profile of contaminations and risks. The median, 25^th^ and 75^th^ quartiles of the seven day simulations were recorded for each of the 50,000 simulations and the average values of the median, 25^th^ and 75^th^ quartiles are reported (Figure 2). A sensitivity analysis was performed to investigate how the variability and uncertainty of the parameters in the model influenced the estimated risks. The sensitivity was estimated using the Spearman’s correlation coefficients between the inputs and outputs of the model. A detailed description of the model and model parameters are found in the Supporting Information (Figure S1, Table S1).

## Results and discussion

### Risks from contaminated surfaces

Risks of SARS-CoV-2 infection from contact with a fomite in community settings are estimated to be low (Figure 1) and influenced by both infection prevalence rate in the community and the frequency with which the fomite is contacted (Figure 2). Median risk of infection from interaction with a contaminated fomite is linearly related to surface contamination, ranging from 2×10^−8^ for a surface with 0.01 RNA genome copies (gc) cm^-2^ to approximately 1 for a surface with ≥10^6^ RNA gc cm^-2^ (Figure 1). Previous studies of surface contamination on public spaces have detected 0.1 to 102 SARS-CoV-2 gc cm^-2 13,15^. In the two studies only 3 of 1281 (0.2%) surfaces sampled were associated with risks of infection greater than 1 in 10,000. The average risk of infection for the sampled surfaces was of 8.5×10^−7^, assuming negligible risks for samples with SARS-CoV-2 RNA below the LOD (1203 out of 1281 surfaces).

**Figure 1.**
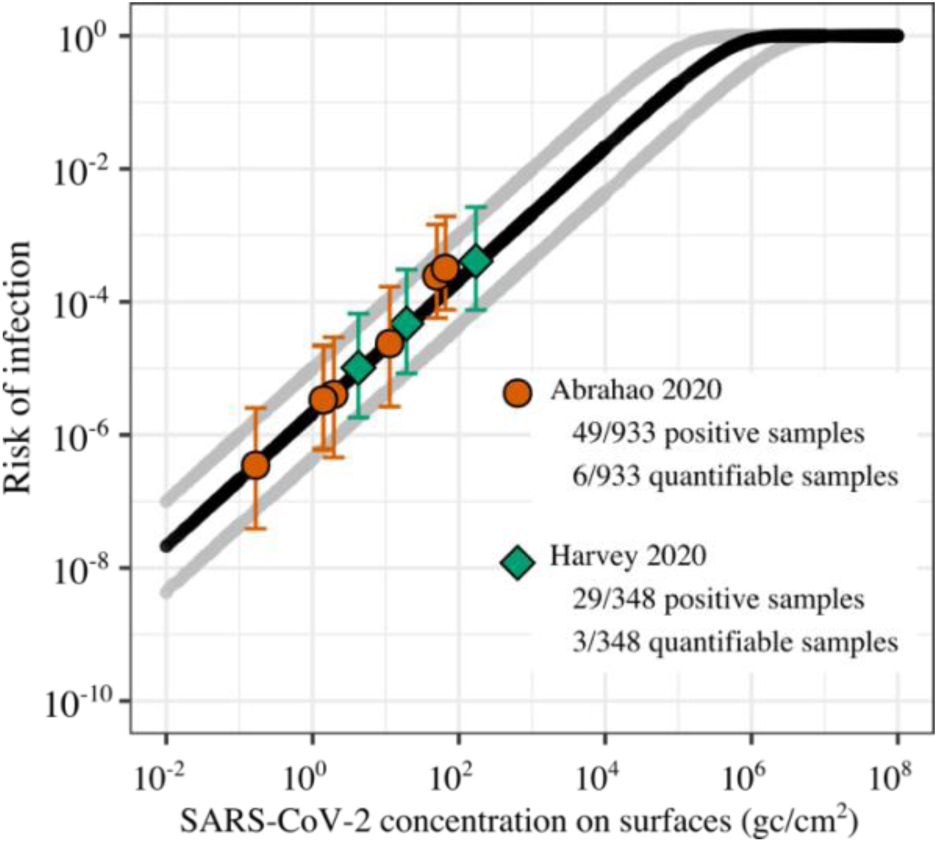
Risk of SARS-CoV-2 infection (unitless, from 0 to 1) as a function of virus concentration on surfaces (genome copies (gc)/cm^2^). Median risk of infection is shown in a continuous black line; Gray lines display the 5^th^ and 95^th^ percentiles. Orange circles^13^ and green diamonds^15^ represent the median risk estimates for point values of surface contamination in public spaces with whiskers from the 5^th^ to the 95^th^ percentiles. Data from Abrahao et al., orange circles, shows the risk for the 6 quantifiable samples out of the 49 RNA positive samples. Data from Harvey et al., green diamonds, shows the risk of 3 quantifiable samples out of the 29 RNA positive samples.

**Figure 3.**
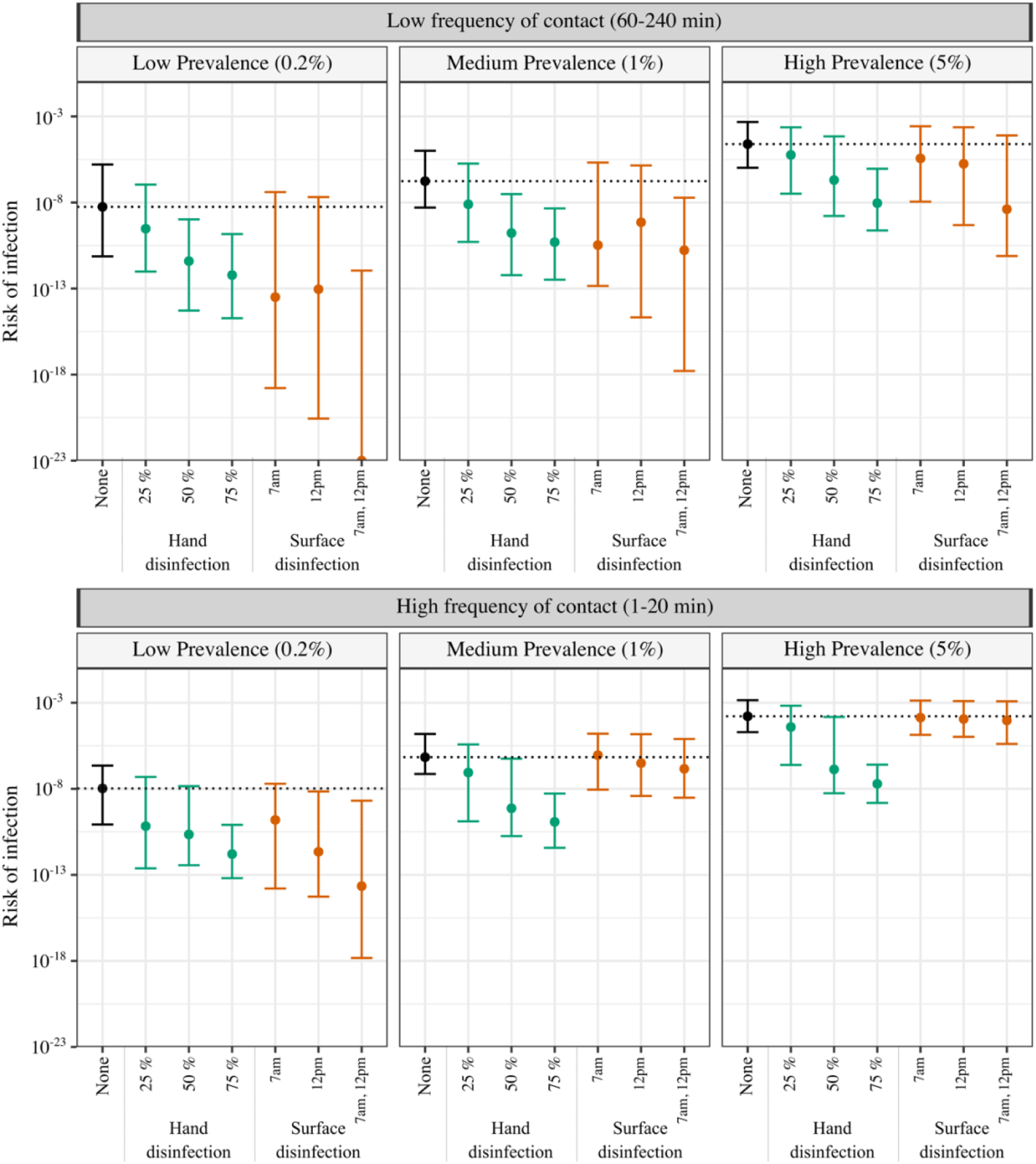
Predicted community-based risk of SARS-CoV-2 infection due to hand-to-surface followed by hand-to-face contact. The plot shows the average median risk of infection, with whiskers from the 25^th^ to the 75^th^ percentiles. Two interventions were tested (hand disinfection [green] and surface disinfection [orange]) in parallel to no intervention control [black]. Compliance for hand disinfection was set to 25, 50, and 75% of the population. Surface disinfection regimes were: every day at 7am, 12pm, or 7am and 12pm. The horizontal black dotted line illustrates the median risk of infection without intervention. Two contact frequencies and three prevalence levels (percentage of the population sick at any given time) were modeled: high contact frequency [1-20 min] and low contact frequency [60-240 min] and low [0.2%], medium [1%], and high [5%] prevalence. The risk of infection of 10^−6^ is equivalent to one person sick as a result of hand-to-mouth contact every million people touching the surface.

### Risks from surface-mediated community transmission

When modeling risks of surface contamination within communities, average median value [IQR] risks range from approximately 1.6×10^−4^ [2.0×10^−5^, 1.4×10^−3^] for the highest risk scenario (5% infection prevalence rate, object contacted once every 1-20 minutes) to 5.6×10^−9^ [7.4×10^−12^, 1.6×10^−6^] in the lowest risk scenario (0.2% prevalence rate, object contacted once every 1-4 hours) (Figure 2). The overwhelming majority of interactions with fomites modeled were associated with risks < 10^−4^ (Table S2). The low risks of community transmission of SARS-CoV-2 via fomites is in accordance with previous studies and opinions of fomite-mediated transmission in hospitals^4–7^.

According to the sensitivity analysis, the model parameters most influencing estimated infection risks within a community are transfer efficiency between the surface and the hand, *TE*_*sh*_, and concentration of SARS-CoV-2 in sputum or saliva, *C*_*sp*_ (Table S1, Figure S4). *TE*_*sh*_ was inversely correlated with risk (Spearman’s rank correlation, ρ = −0.58) and *C*_*sp*_ was directly correlated (ρ = 0.29). Correlation was low with all other modeled parameters (ρ < 0.05).

### Effectiveness of hand and surface disinfection

Hand hygiene was consistently the most effective intervention. Alcohol-based hand disinfectants are portable, widely available, and effective at inactivating coronavirus ^54,55^. Even with low compliance, representative of only 1 in 4 people disinfecting hands after surface contact, median infection risks from fomite contact were reduced by 0.6-2.2 log_10_. Under high compliance, representing 3 of every 4 people disinfecting, median risks decreased by 3.8-4.3 log_10_. Importantly, the impact of hand hygiene also appears to be independent of surface contact frequency and prevalence rates, suggesting a strategy of hand disinfection promotion in community settings is universally applicable. Our findings re-affirm the existing strategies of promoting hand hygiene and making hand disinfect products widely available in shared community settings^56^.

Although the risks of SARS-CoV-2 transmission via fomites are estimated to be low, they are possible and may contribute a small number of new cases during outbreaks. For both surfaces with quantified contamination and modeled surfaces within a community, infection risk estimates are very low when people interact with a single fomite. However, a person’s infection risk increases when accounting for the hundreds of objects contacted every hour, and the thousands of frequently contacted objects (crosswalk buttons, public transportation buttons, ATMs, and railings) within a city. Each interaction provides an opportunity for SARS-CoV-2 transmission. Risk of infection from multiple contacts with fomites – as compared to a single contact with a fomite – is substantially higher. Nevertheless, in our models the risk of infection from a fomite is orders of magnitude lower than the prevalence rates, suggesting the relative contribution of fomite-mediated transmission might be small compared to other transmission routes.

The data used to quantify risks from measured concentrations of SARS-CoV-2 RNA on surfaces in public spaces were obtained from two locations: Somerville, Massachusetts, USA^15^, and Belo Horizonte, Minas Gerais State, Brazil^13^. The sampling collection for both studies occurred throughout a COVID-19 outbreak from March-June 2020. Both places had control measures when the collection took place, including mandatory use of masks in public spaces. The mask use requirement may have influenced surface contamination, with the measured SARS-CoV-2 RNA concentrations lower than what could be observed without a mask requirement. Our modeled interventions included hand disinfection and surface disinfection, but given the widespread use of masks within a community, masks may also help to curb fomite-mediated transmission. Masks are repeatedly shown to be effective at reducing transmission of SARS-CoV-2^57^ through the proposed mechanism of limiting both production of and exposure to aerosolized droplets. Masks may also influence fomite-mediated transmission by reducing hand or surface contamination from droplets and/or reducing hand-to-mouth contact frequency. As there is currently insufficient data on the effectiveness of masks against droplet production and on the frequency of hand-to-mouth contacts, mask use could not be considered as an intervention here.

The model findings are influenced by the model implementation and assumptions, and changes in assumptions may shift some of our conclusions. First, absolute infection risks from QMRA may be unreliable due to the uncertainty and/or variability in the estimates of the parameters^58^. The exponential dose-response model in particular suffers from a number of limitations: the model is based on data of SARS-CoV and Murine hepatitis virus (MHV-1) infection in mice by intranasal administration^40,41^. Extrapolating the model from mice to people and from MHV-1 and SARS-CoV to SARS-CoV-2, introduces uncertainty in infection risk estimates, but – in accordance with current best practice ^59^ – we did not consider this here. Nevertheless, dose-response relationships derived from animal studies tend to be more conservative^60^. An additional limitation is that the dose-response relationship was determined using virus as measured in units of Plaque Forming Units (PFU) and therefore a ratio of genome copies to PFU is needed. The assumed range of ratios of 1:100 to 1:1000 for genome copies to viable virus is based on Influenza, along with the sparse data currently available for SARS-CoV-2. Data quantifying viable virus on fomites in communities would be the “gold standard”, but detection of viable virus is unlikely given previously observed concentrations of SARS-CoV-2 RNA align with estimates of viable virus of <1 / 100 cm^2^. Because of the uncertainties in parameter estimates, QMRA estimates of relative risk reduction from interventions are viewed as more reliable because potential biases in data are incorporated into both the intervention and control risk estimates^58^.

Additional model characteristics likely influence risk estimates. Model parameters used for virus transfer and decay rates are determined experimentally in laboratory conditions and could be different in environmental conditions. Also, prevalence rates modeled here are assumed to correspond directly with the percent of people who are infected and contact the surface with a hand contaminated by coughing. In reality, an unknown fraction of infected people would likely either: 1) stay at home (i.e., quarantine and/or isolation), or 2) not cough directly on their hand. In this regard, the modeled infection risks are likely higher (more conservative) than would be expected at the stated community infection prevalence rates.

Despite the limitations of the underlying model, Quantitative Microbial Risk Assessment remains a valuable tool to understand and characterize risks of surface-mediated transmission of SARS-CoV-2 within communities and test the effectiveness of different interventions. Epidemiological investigations and/or structured experimental designs (i.e., randomized controlled trials) are infeasible given that fomite-mediated transmission is likely a rare event and is difficult to decouple from other – more likely – transmission routes. The results presented here add to the evidence supporting the relatively low contribution of fomites in the transmission of SARS-CoV-2^15^, and can inform guidance on potential intervention strategies.

## Data Availability

All the data, models, and analyses are available at GitHub.

https://github.com/pitolana/QMRA_SARS-CoV-2_Surfaces

## Acknowledgements

We thank Danilo Cuccato, Emmanuel Froustey for their inputs on the model, and Diego Marcos, Sunil K. Dogga, Gabriele Micali, Esther Greenwood, Sital Uprety and Elyse Stachler for reviewing the manuscript. A.K.P. was supported by Swiss National Science Foundation – SNSF.

## Author contributions

T.R.J. and A.K.P designed the study. A.K.P. performed the modeling. T.R.J. and A.K.P wrote the manuscript.

## Table of Contents (TOC)

For Table of Contents Only

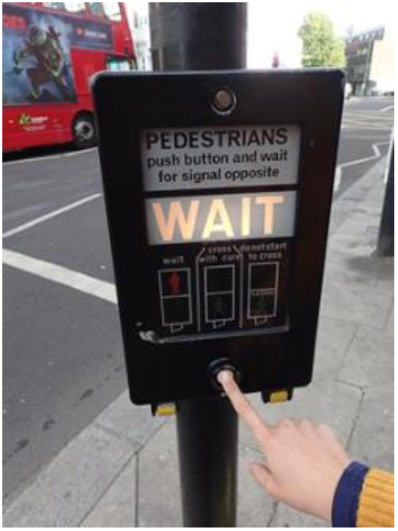

## References

1. Centers for Disease Control (CDC). Scientic Brief: SARS-CoV-2 and Potential Airborne Transmission. 4–7 (2020).

2. World Health Organization. Transmission of SARS-CoV-2: implications for infection prevention precautions. Scientific brief. 1–10 (2020).

3. Ferretti, L. et al. Quantifying SARS-CoV-2 transmission suggests epidemic control with digital contact tracing. Science (80-.). 368, 0–8 (2020).

4. Goldman, E. Exaggerated risk of transmission of COVID-19 by fomites. Lancet Infect. Dis. 20, 892–893 (2020).

5. Mondelli, M. U., Colaneri, M., Seminari, E. M., Baldanti, F. & Bruno, R. Low risk of SARS-CoV-2 transmission by fomites in real-life conditions. Lancet Infect. Dis. 3099, 30678 (2020).

6. Colaneri, M. et al. Lack of SARS-CoV-2 RNA environmental contamination in a tertiary referral hospital for infectious diseases in Northern Italy. J. Hosp. Infect. 105, 474–476 (2020).

7. Colaneri, M. et al. Severe acute respiratory syndrome coronavirus 2 RNA contamination of inanimate surfaces and virus viability in a health care emergency unit. Clin. Microbiol. Infect. 26, 1094.e1–1094.e5 (2020).

8. Boone, S. A. & Gerba, C. P. Significance of fomites in the spread of respiratory and enteric viral disease. Appl. Environ. Microbiol. 73, 1687–1696 (2007).

9. Kraay, A. N. M. et al. Fomite-mediated transmission as a sufficient pathway: a comparative analysis across three viral pathogens. BMC Infect. Dis. (2018). doi:10.1186/s12879-018-3425-x

10. Xiao, S., Li, Y., Wong, T. wai & Hui, D. S. C. Role of fomites in SARS transmission during the largest hospital outbreak in Hong Kong. PLoS One (2017). doi:10.1371/journal.pone.0181558

11. Ong, S. W. X. et al. Air, Surface Environmental, and Personal Protective Equipment Contamination by Severe Acute Respiratory Syndrome Coronavirus 2 (SARS-CoV-2) from a Symptomatic Patient. JAMA - J. Am. Med. Assoc. 2–4 (2020). doi:10.1001/jama.2020.3227

12. Ye, G. et al. Environmental contamination of the SARS-CoV-2 in healthcare premises: An urgent call for protection for healthcare workers. medRxiv Prepr. 1–20 (2020). doi:10.1101/2020.03.11.20034546

13. Abrahao, J. S. et al. Detection of SARS-CoV-2 RNA on public surfaces in a densely populated urban area ofBrazil: A potential tool for monitoring the circulation of infected patients. Sci. Total Environ. (2020). doi:10.1016/j.scitotenv.2020.142645

14. Chia, P. Y. et al. Detection of air and surface contamination by SARS-CoV-2 in hospital rooms of infected patients. Nat. Commun. 11, (2020).

15. Harvey, A. et al. Longitudinal monitoring of SARS-CoV-2 RNA on high-touch surfaces in a community setting. Submitted (2020).

16. Riddell, S., Goldie, S., Hill, A., Eagles, D. & Drew, T. W. The effect of temperature on persistence of SARS-CoV-2 on common surfaces. Virol. J. 17, 145 (2020).

17. Chin, A. et al. Stability of SARS-CoV-2 in different environmental conditions. Lancet Microbe 5247, 2020.03.15.20036673 (2020).

18. van Doremalen, N. et al. Aerosol and Surface Stability of SARS-CoV-2 as Compared with SARS-CoV-1. N. Engl. J. Med. 1–3 (2020). doi:10.1056/NEJMc2004973

19. Julian, T. R., Leckie, J. O. & Boehm, A. B. Virus transfer between fingerpads and fomites. J. Appl. Microbiol. 109, 1868–1874 (2010).

20. Lopez, G. U. et al. Transfer efficiency of bacteria and viruses from porous and nonporous fomites to fingers under different relative humidity conditions. Appl. Environ. Microbiol. 79, 5728–5734 (2013).

21. Rusin, P., Maxwell, S. & Gerba, C. Comparative surface-to-hand and fingertip-to-mouth transfer efficiency of gram-positive bacteria, gram-negative bacteria, and phage. J. Appl. Microbiol. 93, 585–592 (2002).

22. Pitol, A. K., Bischel, H., Kohn, T. & Julian, T. R. Virus transfer at the skin-liquid interface. Environ. Sci. Technol. 51, 14417–14425 (2017).

23. Lu, C. wei, Liu, X. fen & Jia, Z. fang. 2019-nCoV transmission through the ocular surface must not be ignored. The Lancet (2020). doi:10.1016/S0140-6736(20)30313-5

24. Nicas, M. & Best, D. A study quantifying the hand-to-face contact rate and its potential application to predicting respiratory tract infection. J. Occup. Environ. Hyg. 5, 347–352 (2008).

25. Kwok, Y. L. A., Gralton, J. & McLaws, M. L. Face touching: A frequent habit that has implications for hand hygiene. Am. J. Infect. Control 43, 112–114 (2015).

26. Lewis, R. C., Rauschenberger, R. & Kalmes, R. Hand-to-mouth and other hand-to-face touching behavior in a quasi-naturalistic study under controlled conditions ABSTRACT. J. Toxicol. Environ. Heal. Part A 00, 1–7 (2020).

27. Julian, T. R., Canales, R. a., Leckie, J. O. & Boehm, A. B. A model of exposure to rotavirus from nondietary ingestion iterated by simulated intermittent contacts. Risk Anal. 29, 617–632 (2009).

28. Bischel, H. N., Caduff, L., Schindelholz, S., Kohn, T. & Julian, T. R. Health Risks for Sanitation Service Workers along a Container-Based Urine Collection System and Resource Recovery Value Chain. Environ. Sci. Technol. 53, 7055–7067 (2019).

29. Canales, R. A. et al. Modeling the role of fomites in a norovirus outbreak. J. Occup. Environ. Hyg. 16, 16–26 (2019).

30. Adhikari, U. et al. A Case Study Evaluating the Risk of Infection from Middle Eastern Respiratory Syndrome Coronavirus (MERS-CoV) in a Hospital Setting Through Bioaerosols. Risk Anal. 39, 2608–2624 (2019).

31. King, M.-F. et al. Modelling the risk of SARS-CoV-2 infection through PPE doffing in a hospital environment. medRxiv 2020.09.20.20197368 (2020). doi:10.1101/2020.09.20.20197368

32. Jones, R. M. Relative contributions of transmission routes for COVID-19 among healthcare personnel providing patient care. J. Occup. Environ. Hyg. 0, 1–8 (2020).

33. Wilson, A. M. et al. COVID-19 and use of non-traditional masks: how do various materials compare in reducing the risk of infection for mask wearers? J. Hosp. Infect. (2020).

34. Bullard, J. et al. Predicting Infectious Severe Acute Respiratory Syndrome Coronavirus 2 From Diagnostic Samples. Clin. Infect. Dis. 1–4 (2020). doi:10.1093/cid/ciaa638

35. Kim, J. Y. et al. Viral load kinetics of SARS-CoV-2 infection in first two patients in Korea. J. Korean Med. Sci. 35, 1–7 (2020).

36. Ip, D. K. M. et al. The Dynamic Relationship between Clinical Symptomatology and Viral Shedding in Naturally Acquired Seasonal and Pandemic Influenza Virus Infections. Clin. Infect. Dis. 62, 431–437 (2015).

37. ATCC. Converting TCID50 to plaque forming units PFU-124. 1 (2012). Available at: https://www.lgcstandards-atcc.org/Global/FAQs/4/8/Converting_TCID50_to_plaque_forming_units_PFU-124.aspx?geo_country=gb#. (Accessed: 20th October 2020)

38. Wilson, A. M. et al. Evaluating a transfer gradient assumption in a fomite-mediated microbial transmission model using an experimental and Bayesian approach. J. R. Soc. Interface 17, (2020).

39. Watanabe, T., Bartrand, T. A., Weir, M. H., Omura, T. & Haas, C. N. Development of a Dose-Response Model for SARS Coronavirus. 30, (2010).

40. DeDiego, M. L. et al. Pathogenicity of severe acute respiratory coronavirus deletion mutants in hACE-2 transgenic mice. Virology (2008). doi:10.1016/j.virol.2008.03.005

41. De Albuquerque, N. et al. MurineHepatitis Virus Strain 1 Produces a Clinically Relevant Model of Severe Acute Respiratory Syndrome in A/J Mice. J. Virol. (2006). doi:10.1128/jvi.00747-06

42. Zhou, B. et al. SARS-CoV-2 spike D614G variant confers enhanced replication and transmissibility. bioRxiv (2020). doi:https://doi.org/10.1101/2020.10.27.357558

43. Wölfel, R. et al. Virological assessment of hospitalized patients with COVID-2019. Nature 1–14 (2020). doi:10.1038/s41586-020-2196-x

44. Pan, Y., Zhang, D., Yang, P., Poon, L. L. M. & Wang, Q. Viral load of SARS-CoV-2 in clinical samples. Lancet Infect. Dis. 20, 411–412 (2020).

45. To, K. K. W. et al. Temporal profiles of viral load in posterior oropharyngeal saliva samples and serum antibody responses during infection by SARS-CoV-2: an observational cohort study. Lancet Infect. Dis. 20, 565–574 (2020).

46. Bendavid, E. et al. COVID-19 Antibody Seroprevalence in Santa Clara County, California. medRxiv 2020.04.14.20062463 (2020). doi:10.1101/2020.04.14.20062463

47. Perez-Saez, J. et al. Serology-informed estimates of SARS-CoV-2 infection fatality risk in Geneva, Switzerland. Lancet Infect. Dis. 3099, 2–3 (2020).

48. Pollán, M. et al. Prevalence of SARS-CoV-2 in Spain (ENE-COVID): a nationwide, population-based seroepidemiological study. Lancet 396, 535–544 (2020).

49. Erikstrup, C. et al. Estimation of SARS-CoV-2 infection fatality rate by real-time antibody screening of blood donors. Clin. Infect. Dis. (2020). doi:10.1093/cid/ciaa849

50. Amorim Filho, L. et al. Seroprevalence of anti-SARS-CoV-2 among blood donors in Rio de Janeiro, Brazil. Rev. Saude Publica 54, 69 (2020).

51. Nicas, M. & Sun, G. An integrated model of infection risk in a health-care environment. Risk Anal. 26, 1085–1096 (2006).

52. van Doremalen, N. et al. Aerosol and Surface Stability of SARS-CoV-2 as Compared with SARS-CoV-1. N. Engl. J. Med. (2020). doi:10.1056/NEJMc2004973

53. Narendra Kumar Chaudhary et al. Fighting the SARS CoV-2 (COVID-19) Pandemic with Soap. Preprints 060, 1–19 (2020).

54. Rabenau, H. F., Kampf, G., Cinatl, J. & Doerr, H. W. Efficacy of various disinfectants against SARS coronavirus. J. Hosp. Infect. 61, 107–111 (2005).

55. Golin, A. P., Choi, D. & Ghahary, A. Hand sanitizers: A review of ingredients, mechanisms of action, modes of delivery, and efficacy against coronaviruses. Am. J. Infect. Control (2020). doi:10.1016/j.ajic.2020.06.182

56. World Health Organization (WHO). Recommendation to Member States to improve hand hygiene practices widely to help prevent the transmission of the COVID-19 virus. Interim guidance (2020).

57. Liang, M. et al. Efficacy of face mask in preventing respiratory virus transmission: A systematic review and meta-analysis. Travel Med. Infect. Dis. 101751 (2020). doi:10.1016/j.tmaid.2020.101751

58. World Health Organization (WHO). Quantitative microbial risk assessment. Application for water safety management. (2016).

59. Haas, C. N., Rose, J. B. & Gerba, C. P. Quantitative Microbial Risk Assessment: Second Edition. John Wiley & Sons, Inc. (2014). doi:10.1002/9781118910030

60. Haas, C. WikiQMRA: Completed Dose Response Models. Available at: http://qmrawiki.org/framework/dose-response/experiments. (Accessed: 1st October 2020)

