## Supplemental Material for "Community Transmission of SARS-CoV-2 by Fomites: Risks and Risk Reduction Strategies"

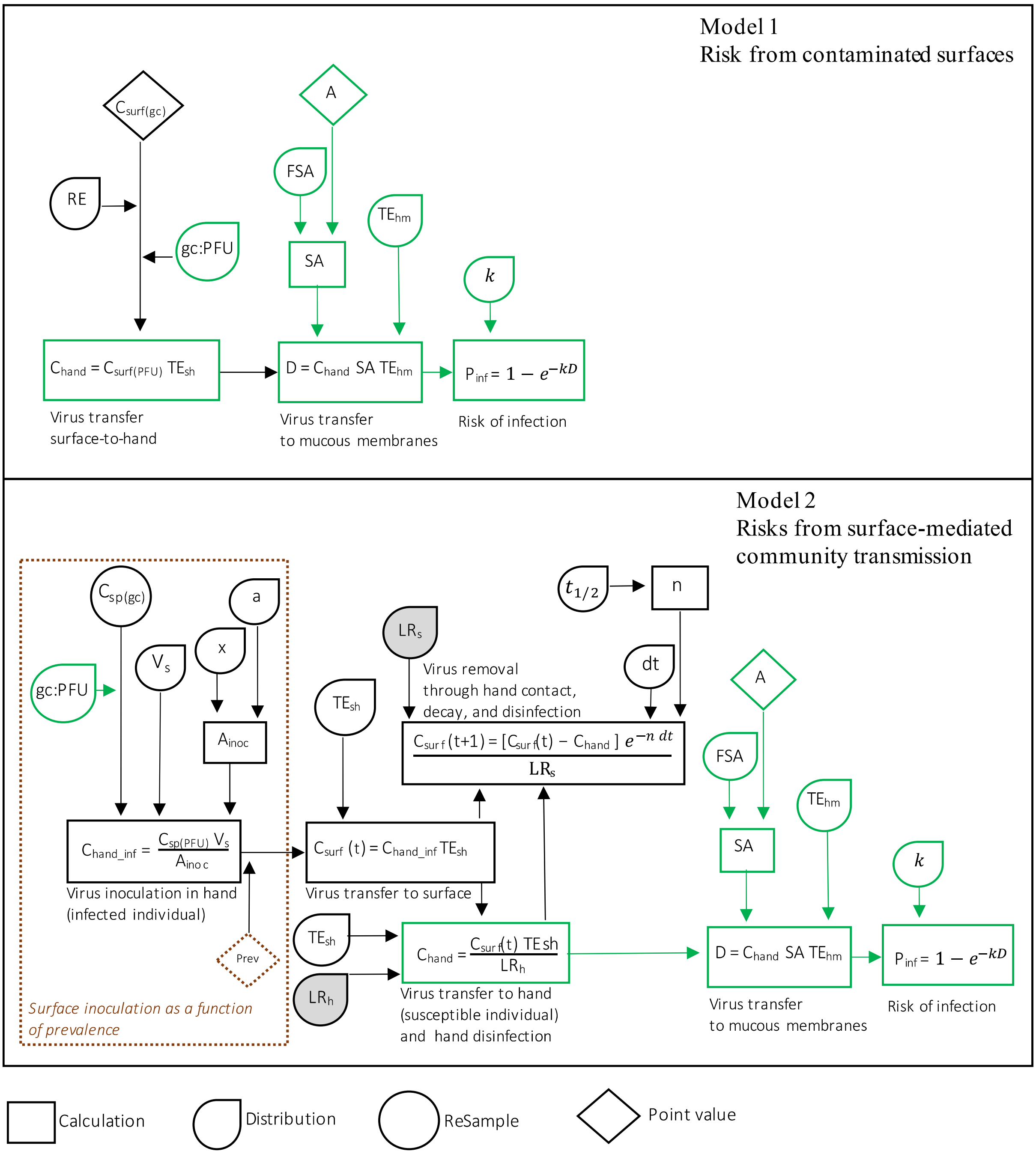


**Figure S1. Diagram of the mathematical models used to estimate the risk from contaminated surfaces and the risk from surface-mediated community-transmission.** Green rectangles represent shared equations or parameters between the risk model for contaminated surface and the risk model for surface-mediated community transmission. Within the dotted brown line we show the model used to calculate the surface contamination as a function of prevalence. Filled shapes represent the parameters only used in the intervention scenarios ($\log_{10}$ reduction values). The descriptions of parameters' and inputs can be found in Table 1.

Table S1. Input parameters for the risk assessment model

| **Parameter** | **Units** | **Description** | **Distributions (Input values)/ Equations** | **Reference and comments** |
|---|---|---|---|---|
| $C_{sp\,(gc)}$ | Gene copies (gc) $mL^{-1}$ | Concentration of SARS-CoV-2 in sputum or saliva | ReSample (data set) | [4–7] RT-qPCR data on viral loads of 9, 2, 2, and 23 patients with COVID-19 |
| $C_{surf\,(gc)}$ | gc $cm^{-2}$ | Concentration of SARS-CoV-2 in surfaces | Point values (102.4, 2.5, 11.6, 6.8, 1.2, 30, 39.3, 0.8, 0.8, 0.1) | [8,9] RT-qPCR data on concentration of SARS-CoV-2 in surfaces found in public spaces |
| gc: PFU | unitless | Genome copies to infectious virus conversion factor. Used to convert the viral concentrations $C_{(gc)}$ to $C_{(PFU)}$ | Uni ($10^2$-$10^3$) | [10,11] Based on the ratio for influenza A(H1N1), A(H3N2), and influenza B and the ratio of $TCID_{50}$ to PFU |
| $RE$ | unitless | Recovery efficiency | Point value (0.6) | [9] Recovery efficiency from swabs |
| $V_s$ | mL | Volume of saliva expelled per cough | Uni (0.0396 - 0.0484) | [12] Volume of 0.044mL<br>[13] Assumed uniform distribution using volume ± 10% |
| $x$ | cm | Distance between hand and mouth | Uni (5-10) | Assumed |
| $a$ | degrees | Right angle of cone | Uni (27.5-35) | Based on the images of people spreading particles while coughing[2,3] |
| $A_{inoc}$ | $cm^2$ | Area of inoculation by cough | $A_{inoc} = \pi r^2$<br>$r = x \tan(a)$ | [1] Calculated assuming viral particles spread conically (Supplementary Figure 1) |
| $C_{hand\_inf}$ | PFU $cm^{-2}$ | Concentration of SARS-CoV-2 on the hands of an infected individual | $\frac{C_{sp}\,V_s}{A_{inoc}}$ | [1] Calculated assuming viral particles spread conically (Supplementary Figure 1) |
| $t_{\frac{1}{2}stl}$ | min | Half-life of SARS-CoV-2 in metal | N (338,35) | [14]Based on SARS-CoV-2 infectivity at 40% RH and 21-23°C |
| $t_{\frac{1}{2}pl}$ | min | Half-life of SARS-CoV-2 in plastic | N (409,39) | [14]Based on SARS-CoV-2 infectivity at 40% RH and 21-23°C |
| $n$ | $min^{-1}$ | Exponential decay constant | $\frac{\ln 2}{t_{1/2}}$ | Calculated assuming exponential decay |
| dt | min | Time between surface touching | Uni (1-20)<br>Uni (60-240) | Based on public transport schedules in major cities<br>Contact with surfaces was assumed to happen between 7 am and 11pm |
| $TE_{hm}$ | % | Transfer efficiency from hand to mucous membranes | N (20,6.3) | [15]Transfer efficiency of viruses (MS2) from hand to saliva |
| $TE_{sh\_stl}$ | % | Transfer efficiency of virus between metal and hand | N (37.4, 16) | [16]Transfer efficiency of viruses (MS2) between steel and hand at 40-65% RH |
| $TE_{sh\_pl}$ | % | Transfer efficiency of virus between plastic and hand | N (79.5,21.2) | [16]Transfer efficiency of viruses (MS2) between plastic and hand at 40-65% RH |

| | | | | |
|---|---|---|---|---|
| $LR_s$ | | $Log_{10}$ reduction for surface disinfection | Uni (3-4) | [17,18] $Log_{10}$ reduction of coronaviruses on surfaces with ethanol and chlorine disinfection |
| $LR_h$ | | $Log_{10}$ reduction for hand disinfection | Point value (4.25) | [19] $Log_{10}$ reduction of SARS-CoV with alcohol-based (>75%) sanitizer |
| $Prev$ | % | Prevalence | Point values<br>Low (0.2%)<br>Medium (1%)<br>High (5%) | Medium prevalence based on rates encountered during the peak of the first wave of COVID-19 in major cities[20–24]. |
| $C_{surf(t+1)}$ | PFU $cm^{-2}$ | Concentration of SARS-CoV-2 in surface at time = t+1 | $\frac{(C_{surf(t)}-C_{hand})e^{-n\,dt}}{LR_h}$ | Calculated |
| $C_{hand}$ | PFU $cm^{-2}$ | Concentration of SARS-CoV-2 on the hands of susceptible individuals | $\frac{C_{surf}\,TE_{sh}}{LR_h}$ | Calculated |
| $SA$ | $cm^2$ | Surface area in contact with mucous membranes | Uni (3.9-5.9) | [25] Fractional surface area for partial finger. [26]Average hand surface area |
| $D$ | PFU | Dose | $C_{hand}\,SA\ TE_{sh}$ | Calculated |
| $k$ | $PFU^{-1}$ | Parameter of exponential dose-response | Tri (0.00107, 0.00135, 0.00680) | [27] Data obtained from QMRAwiki, based on 2 studies [28] [29] using the $0.5^{th}$, $50^{th}$, and $99.5^{th}$ percentiles as min, mode, and max |
| $P_{inf}$ | unitless | Probability of infection | $1-e^{-kD}$ | Calculated<br>[27] Model obtained from QMRAwiki |

Distributions and input parameters are abbreviated as follows: N= Normal (mean, SD), Uni =Uniform (min-max), Tri=Triangular (min, mode, max). ReSample refers to random sampling with replacement from the data set of of viral loads reported for patients with COVID-19 in the associated reference.

**Inoculation of viruses on hands**

We assumed that a cough spread particles conically[1] (Figure 1). The concentration of viruses (virus $cm^{-2}$) was estimated by calculating the surface area of a circle projected by the cone at a distance $x$ from the mouth (Equations 1 and 2). Distance $x$ was assumed to be a uniform distribution between 5 and 10 cm. The conical opening angle, between 27.5-35°, was informed by images of people spreading particles while coughing[2,3].

$$r = x \tan(a) \qquad \text{(Equation 1)}$$

$$A_{inoc} = \pi r^2 \qquad \text{(Equation 2)}$$

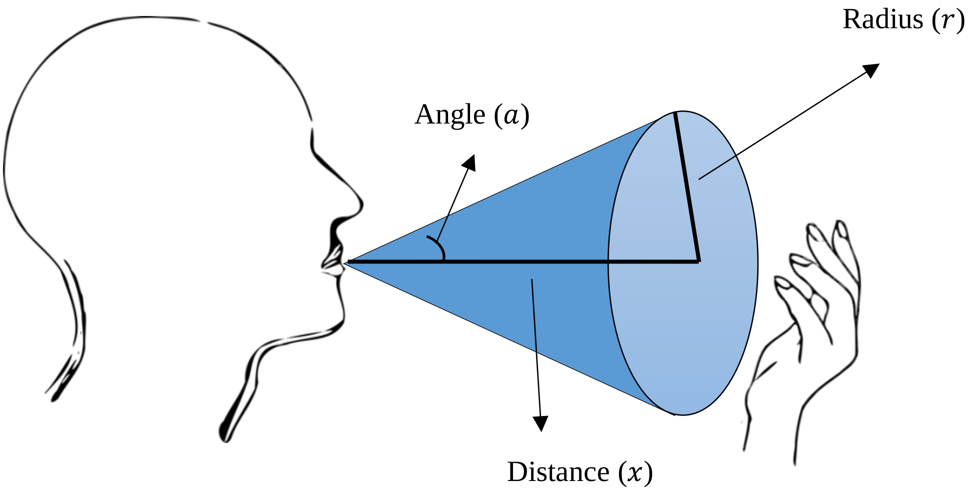


**Figure S2.** Conical distribution of particles through a cough

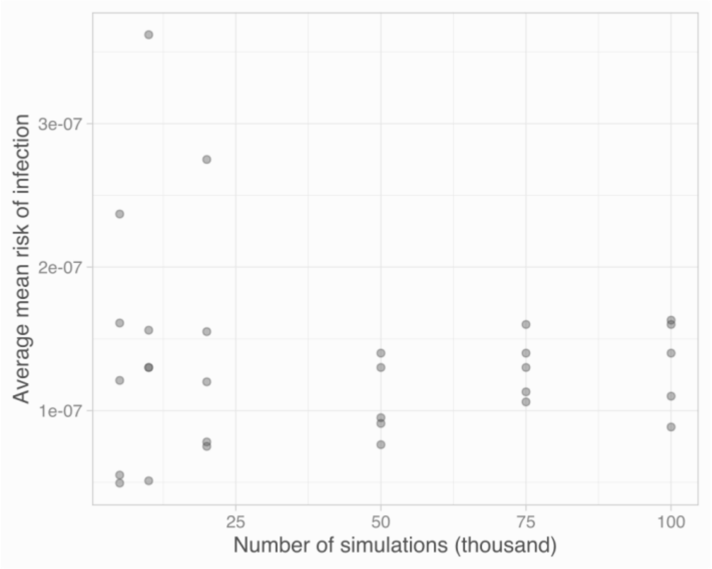


**Figure S3. Risk of infection *vs* number of Monte Carlo simulations.** The analyses were run five times for the baseline scenario (Prevalence 1%, no intervention) for 5000, 10000, 20000, 50000, 75000, and 100000 simulations. The average median risk of infection is shown in black circles.

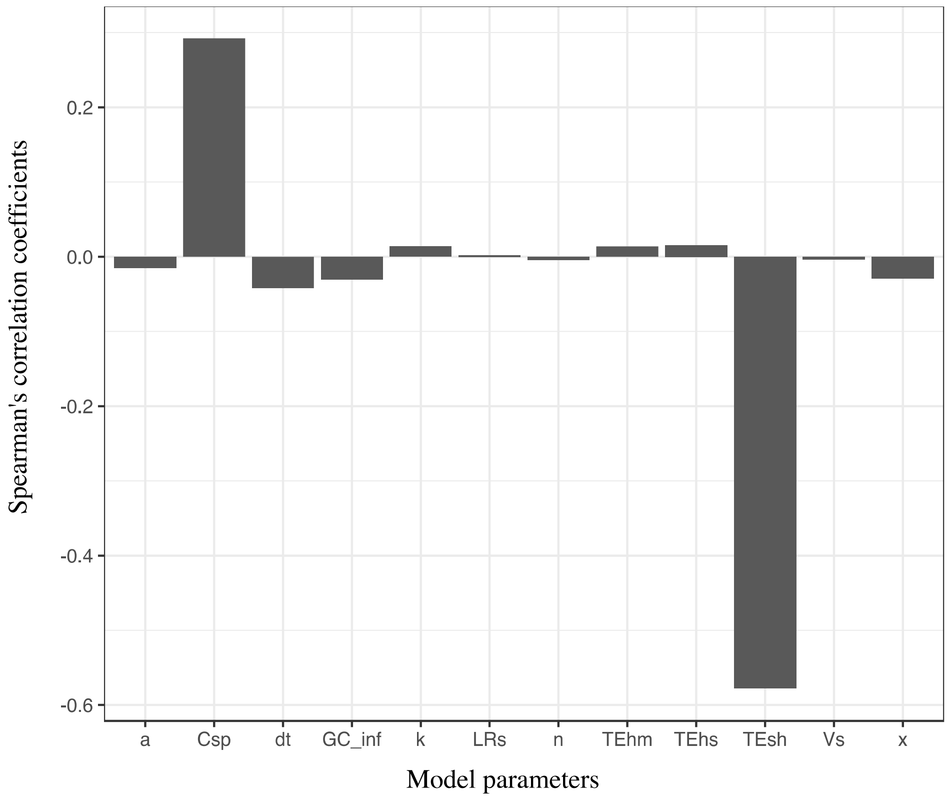


**Figure S3. Sensitivity analysis for the "Risks from surface-mediated community transmission" model.** Spearman's correlation coefficients for the parameters used in the community transmission model. Parameters are abbreviated as follows: a = opening angle of right cone, Csp = Concentration of SARS-CoV-2 in the sputum or saliva of patients, dt = contact frequency, GC_inf = genome copies(gc) per Plaque Forming Units (PFU) ratio, k = parameter of the dose-response exponential model, LRs = $Log_{10}$ reduction due to surface disinfection, n = exponential decay constant, surf_dis = $log_{10}$ reduction for surface disinfection, TEhm = transfer efficiency of viruses from hand to mouth, TEhs = transfer efficiency of viruses from hand to surface, TEsh = transfer efficiency of viruses from surface to hand, Vs = volume of saliva expelled per cough, x= distance between hand and mouth.

**Table S2.** Percentage of Contacts with Estimated Risks Above $10^{-4}$

| | **Percentage of Contacts with Estimated Risks Above 1 in 10,000 (%)** | | | | | |
|---|---|---|---|---|---|---|
| | Low Frequency Contacts | | | High Frequency Contacts | | |
| | Low Prevalence | Medium Prevalence | High Prevalence | Low Prevalence | Medium Prevalence | High Prevalence |
| No Intervention | 0.8 | 3.8 | 17.2 | 1.4 | 6.9 | 27.6 |
| Hand Disinfection (compliance) | | | | | | |
| 25% | 0.6 | 2.7 | 12.9 | 1.1 | 5.2 | 21.0 |
| 50% | 0.4 | 1.9 | 9.3 | 0.8 | 3.4 | 14.8 |
| 75% | 0.2 | 0.7 | 5.5 | 0.3 | 1.7 | 6.8 |
| Surface Disinfection (times a day) | | | | | | |
| Once (7am) | 0.5 | 2.8 | 12.9 | 1.4 | 6.6 | 27.0 |
| Once (12pm) | 0.5 | 2.6 | 12.0 | 1.4 | 6.6 | 26.8 |
| Twice (7am,12pm) | 0.4 | 2.0 | 9.5 | 1.4 | 6.4 | 26.5 |

Low prevalence = 0.2% of the population was assumed to have the disease, medium prevalence = 1% of the population, and high Prevalence = 5% of the population.

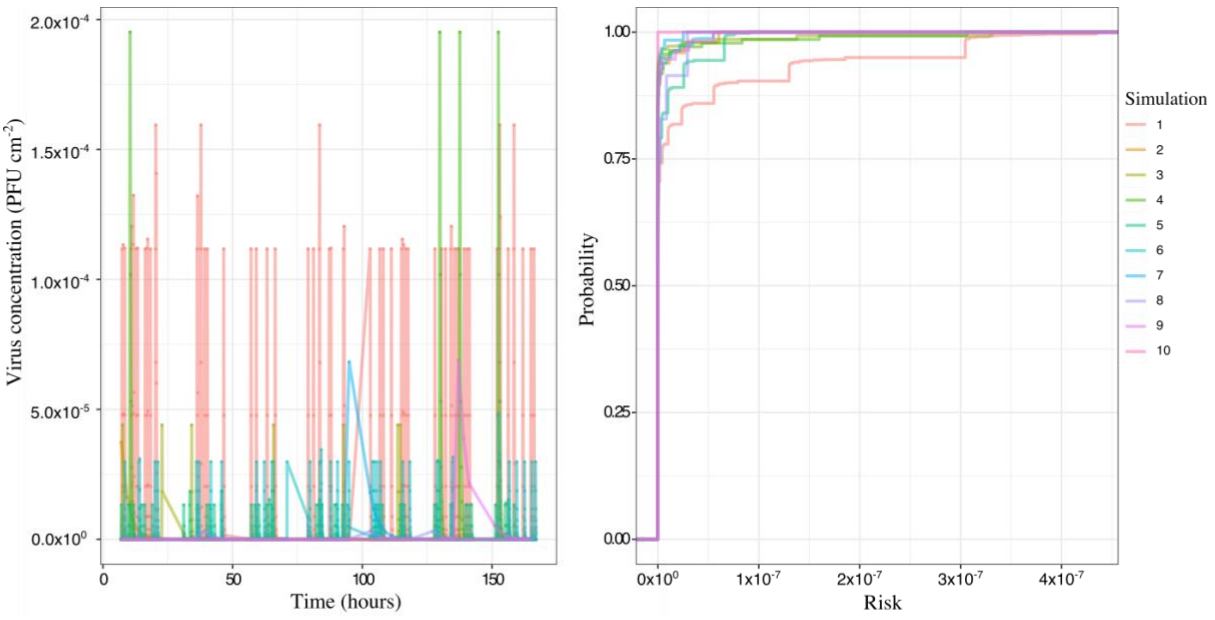


**Figure S4.** Modeled virus concentration across time for ten simulations (left), cumulative distribution function for risks for the same simulations (right). The community transmission model estimates concentrations across time for seven days (168 hours); inoculation of SARS-CoV-2 into the surface is considered to happen between 7 am and 11 pm.

**References**

1. Nicas, M. & Sun, G. An integrated model of infection risk in a health-care environment. *Risk Anal.* **26**, 1085–1096 (2006).
2. Bandiera, L. *et al.* Face Coverings and Respiratory Tract Droplet Dispersion. 1–15 (2020).
3. Bourouiba, L., Dehandschoewercker, E. & Bush, J. W. M. Violent expiratory events: On coughing and sneezing. *J. Fluid Mech.* **745**, 537–563 (2014).
4. Kim, J. Y. *et al.* Viral load kinetics of SARS-CoV-2 infection in first two patients in Korea. *J. Korean Med. Sci.* **35**, 1–7 (2020).
5. Wölfel, R. *et al.* Virological assessment of hospitalized patients with COVID-2019. *Nature* 1–14 (2020). doi:10.1038/s41586-020-2196-x
6. Pan, Y., Zhang, D., Yang, P., Poon, L. L. M. & Wang, Q. Viral load of SARS-CoV-2 in clinical samples. *Lancet Infect. Dis.* **20**, 411–412 (2020).
7. To, K. K. W. *et al.* Temporal profiles of viral load in posterior oropharyngeal saliva samples and serum antibody responses during infection by SARS-CoV-2: an observational cohort study. *Lancet Infect. Dis.* **20**, 565–574 (2020).
8. Abrahao, J. S. *et al.* Detection of SARS-CoV-2 RNA on public surfaces in a densely populated urban area ofBrazil: A potential tool for monitoring the circulation of infected patients. *Sci. Total Environ.* (2020). doi:10.1016/j.scitotenv.2020.142645
9. Harvey, A. *et al.* Longitudinal monitoring of SARS-CoV-2 RNA on high-touch surfaces in a community setting. *Submitted* (2020).
10. Ip, D. K. M. *et al.* The Dynamic Relationship between Clinical Symptomatology and Viral Shedding in Naturally Acquired Seasonal and Pandemic Influenza Virus Infections. *Clin. Infect. Dis.* **62**, 431–437 (2015).
11. ATCC. Converting TCID50 to plaque forming units PFU-124. 1 (2012). Available at: https://www.lgcstandards-atcc.org/Global/FAQs/4/8/Converting_TCID50_to_plaque_forming_units_PFU-124.aspx?geo_country=gb#. (Accessed: 20th October 2020)
12. Nicas, M. & Jones, R. M. Relative contributions of four exposure pathways to influenza

infection risk. *Risk Anal.* **29**, 1292–1303 (2009).

13. Adhikari, U. *et al.* A Case Study Evaluating the Risk of Infection from Middle Eastern Respiratory Syndrome Coronavirus (MERS-CoV) in a Hospital Setting Through Bioaerosols. *Risk Anal.* **39**, 2608–2624 (2019).
14. van Doremalen, N. *et al.* Aerosol and Surface Stability of SARS-CoV-2 as Compared with SARS-CoV-1. *N. Engl. J. Med.* (2020). doi:10.1056/NEJMc2004973
15. Pitol, A. K., Bischel, H., Kohn, T. & Julian, T. R. Virus transfer at the skin-liquid interface. *Environ. Sci. Technol.* **51**, 14417–14425 (2017).
16. Lopez, G. U. *et al.* Transfer efficiency of bacteria and viruses from porous and nonporous fomites to fingers under different relative humidity conditions. *Appl. Environ. Microbiol.* **79**, 5728–5734 (2013).
17. Hulkower, R. L., Casanova, L. M., Rutala, W. A., Weber, D. J. & Sobsey, M. D. Inactivation of surrogate coronaviruses on hard surfaces by health care germicides. *Am. J. Infect. Control* **39**, 401–407 (2011).
18. Sattar, S. A., Springthorpe, V. S., Karim, Y. & Loro, P. Chemical disinfection of non-porous inanimate surfaces experimentally contaminated with four human pathogenic viruses. *Epidemiol. Infect.* **102**, 493–505 (1989).
19. Rabenau, H. F., Kampf, G., Cinatl, J. & Doerr, H. W. Efficacy of various disinfectants against SARS coronavirus. *J. Hosp. Infect.* **61**, 107–111 (2005).
20. Bendavid, E. *et al.* COVID-19 Antibody Seroprevalence in Santa Clara County, California. *medRxiv* 2020.04.14.20062463 (2020). doi:10.1101/2020.04.14.20062463
21. Perez-Saez, J. *et al.* Serology-informed estimates of SARS-CoV-2 infection fatality risk in Geneva, Switzerland. *Lancet Infect. Dis.* **3099**, 2–3 (2020).
22. Pollán, M. *et al.* Prevalence of SARS-CoV-2 in Spain (ENE-COVID): a nationwide, population-based seroepidemiological study. *Lancet* **396**, 535–544 (2020).
23. Erikstrup, C. *et al.* Estimation of SARS-CoV-2 infection fatality rate by real-time antibody screening of blood donors. *Clin. Infect. Dis.* (2020). doi:10.1093/cid/ciaa849
24. Amorim Filho, L. *et al.* Seroprevalence of anti-SARS-CoV-2 among blood donors in Rio de

Janeiro, Brazil. *Rev. Saude Publica* **54**, 69 (2020).

25. AuYeung, W., Canales, R. A. & Leckie, J. O. The fraction of total hand surface area involved in young children’s outdoor hand-to-object contacts. *Environ. Res.* **108**, 294–299 (2008).
26. US Environmental Protection Agency. Exposure Factors Handbook: 2011 Edition. *U.S. Environ. Prot. Agency* **1**, 1–1466 (2011).
27. Haas, C. WikiQMRA: Completed Dose Response Models. Available at: http://qmrawiki.org/framework/dose-response/experiments. (Accessed: 1st October 2020)
28. De Albuquerque, N. *et al.* MurineHepatitis Virus Strain 1 Produces a Clinically Relevant Model of Severe Acute Respiratory Syndrome in A/J Mice. *J. Virol.* (2006). doi:10.1128/jvi.00747-06
29. DeDiego, M. L. *et al.* Pathogenicity of severe acute respiratory coronavirus deletion mutants in hACE-2 transgenic mice. *Virology* (2008). doi:10.1016/j.virol.2008.03.005
